# Clinical validation of an HPV whole genome sequencing assay for molecular residual disease detection in HPV-associated head and neck cancer patients treated with surgery

**DOI:** 10.1101/2025.11.24.25340887

**Authors:** Shun Hirayama, Yana Al-Inaya, Michael E. Bryan, Dipon Das, Ling Aye, Saskia Naegele, Julia Mendel, William C. Faquin, Peter M. Sadow, Matthew G. Crowson, Derrick Lin, Mark Varvares, Allen L. Feng, Daniel Deschler, Jonathan Paly, Ross Merkin, Thomas J. Roberts, Michael Lawrence, A. John Iafrate, Lori Wirth, Adam S. Fisch, Zoe Guan, Jeremy Richmon, Daniel L. Faden

## Abstract

**Background:** Surgery is a common treatment for early-stage HPV-associated head and neck squamous cell carcinoma (HPV+HNC). Selection of patients who require adjuvant treatment is based on clinicopathologic risk factors, which have poor individualized prognostic capacity. Circulating tumor HPV DNA (ctHPVDNA) is a highly sensitive and specific biomarker for HPV+HNC at diagnosis, but current clinically available assays lack the necessary sensitivity for accurate minimal residual disease (MRD) detection after surgery. Here, we applied a significantly more sensitive HPV whole genome sequencing (WGS) assay to determine the prognostic value of ctHPVDNA-based MRD detection and compare this head-to-head with existing approaches and clinical standard of care.

**Patients and methods:** 103 patients with AJCC 8 Stage I-IV HPV+HNC treated with definitive surgery were prospectively enrolled. Blood was collected before surgery, after surgery, and in surveillance and analyzed by clinically validated HPV WGS and droplet digital (dd)PCR assays. The primary hypothesis tested was that patients with MRD detection after surgery would have inferior disease-free survival (DFS) and overall survival (OS).

**Results:** With a median follow up of 27 months, patients with ctHPVDNA detected after surgery had significantly worse 2-year DFS and OS compared to those without ctHPVDNA (DFS 60%, 95% CI:31-80% vs 100%, *p<*0.001; OS 73%, 95% CI:43-89% vs 98%, 95% CI:88-100, *p=*0.002). Patients with ctHPVDNA detected following multi-modality treatment completion also had significantly worse 2-year DFS and OS compared to patients without ctHPVDNA (DFS 0% vs 100%, *p*<0.001) (OS 50%, 95% CI:11-80% vs 100%, *p<*0.001).

MRD status was a stronger predictor of DFS than standard clinicopathologic criteria (HR 25.2; p = 0.003). Lead time from molecular detection of recurrence to clinical detection of recurrence was up to 17.5 months and nearly twice as long compared to ddPCR (7.1 vs 4.1 months).

**Conclusion:** Applying an ultrasensitive HPV WGS liquid biopsy, HPV+HNC patients with ctHPVDNA detected after surgery and following treatment completion had significantly worse DFS and OS, highlighting the potential for MRD status for personalized adjuvant treatment decision-making.

**Highlights:**

- Applying an ultrasensitive HPV WGS liquid biopsy, HPV+HNC patients with MRD after surgery and following treatment completion had significantly worse DFS and OS
- Patients with MRD had a strong benefit from adjuvant treatment in decreasing recurrence and death while patients without MRD showed no benefit
- MRD status after surgery outperformed conventional clinicopathologic features in predicting recurrence
- Median lead time from molecular detection of recurrence to clinical detection was >7 months, and up to 17.5 months, double existing liquid biopsy approaches

## Introduction

Human papillomavirus-associated head and neck squamous cell carcinoma (HPV+HNC) is increasing in incidence^1–4^. The majority of HPV+HNC patients present with early-stage disease and are frequently treated with surgery to remove both the primary tumor and involved lymph nodes^5, 6^. Most patients who undergo surgery receive adjuvant treatment with radiation or chemoradiation^5, 6^. While survival is favorable with this treatment paradigm, treatments carry life-long side effects, significantly impacting quality of life in domains such as swallowing function^7^. There is significant interest in de-escalating treatment by withholding adjuvant treatment to lessen side effects, yet currently, estimation of residual disease after surgery—and consequently, decisions regarding adjuvant therapy—are based on clinicopathologic risk factors, which lack reliable prognostic accuracy on an individual level. There is a critical need for biomarkers capable of accurately detecting minimal residual disease (MRD) in the immediate postoperative period to enable personalized, risk-adapted adjuvant treatment decisions.

HPV+HNCs release circulating tumor HPV DNA (ctHPVDNA), enabling a real-time, tumor-specific biomarker of disease burden^8, 9^. ctHPVDNA detection offers several key advantages over conventional circulating tumor DNA (ctDNA) approaches that rely on somatic mutation profiling, which are being widely utilized in solid tumors types such as lung, colorectal, and HPV-independent head and neck cancers^10–12^. Currently, the most sensitive ctDNA assays are “tumor-informed,” requiring a tissue biopsy for high-depth next-generation sequencing (NGS) to design a custom panel tailored to each patient’s tumor genome^13^. These approaches are inherently limited by their high costs associated with bespoke assay creation, and long turnaround times, which often preclude timely MRD assessment in real-world clinical workflows—particularly for HPV+HNC, where MRD status must be available within 2-3 weeks post-surgery to guide adjuvant treatment initiation within a 6-week window^14, 15^. ctHPVDNA-based approaches overcome these limitations by targeting the HPV genome, which is highly conserved, small in size, shared across patients, lacks homology to human DNA, and can be present in higher copy numbers^16^. As a result, these assays do not require tumor tissue biopsy or bespoke design— significantly reducing both turnaround time and cost. Because of these advantages, droplet digital PCR (ddPCR)-based methods, which target a small portion of the HPV genome, for the most common HPV genotypes, have become widely adopted at diagnosis and during post-treatment surveillance of HPV+HNC^17–22^.

Studies utilizing ddPCR-based ctHPVDNA detection have demonstrated that ctHPVDNA clears rapidly—within hours—following complete surgical resection in HPV+HNC, but remains persistently elevated in patients with macroscopic residual disease^23^. Moreover, multiple retrospective analyses have shown that postoperative ctHPVDNA levels are significantly associated with 2-year overall survival (OS) and disease-free survival (DFS), supporting its potential as a prognostic biomarker for MRD^24–26^.

Although ctHPVDNA holds significant promise as a biomarker for MRD, current ddPCR-based approaches lack the sensitivity required for clinical application. In the first prospective study to evaluate ctHPVDNA as an integral biomarker for treatment de-escalation, Chen et al. found that ctHPVDNA detection was insufficient to guide decisions regarding the omission of adjuvant radiation^27^. Among 12 patients who underwent surgery with negative margins and had two postoperative negative ctHPVDNA tests within six weeks of surgery, 25% experienced radiographic recurrence and ultimately required radiation therapy. Notably, one patient developed biopsy-confirmed gross nodal recurrence despite persistently negative ctHPVDNA results. These findings are further supported by retrospective data from the De-escalated Adjuvant Radiation Therapy (DART) trial, where the 24-month DFS was only 63.6% among ctHPVDNA-negative patients with pN2 disease who received de-escalated radiation^25^. Together, these results underscore the need for more sensitive approaches to accurately guide postoperative treatment decisions through accurate MRD detection.

NGS-based ctHPVDNA analysis platforms have recently emerged, offering markedly improved sensitivity over current ddPCR-based methods by targeting the entire HPV genome (whole-genome sequencing (WGS)) across all HPV genotypes^8, 28, 29^. In a head-to-head comparison, HPV-DeepSeek—a multi-feature WGS-based ctHPVDNA assay—demonstrated significantly higher diagnostic accuracy (99%) than ddPCR (90%) at the time of cancer diagnosis. This advantage was especially pronounced in early-stage disease; for example, in patients with T1 tumors and 0–1 lymph nodes ≤3 cm, HPV-DeepSeek achieved 100% diagnostic accuracy compared to 55% for ddPCR, suggesting that WGS-based assays may overcome the limitations of existing ddPCR-based approaches in settings with ultra-low ctHPVDNA levels, such as MRD.

Here we report findings from a prospective observational trial of 103 patients with HPV+HNC treated with curative-intent surgery. Our primary objective was to evaluate the prognostic and predictive utility of ctHPVDNA-based MRD detection using HPV-DeepSeek and its association with DFS and OS. We also compared the prognostic performance of HPV-DeepSeek with that of ddPCR, assessed the lead time between molecular detection and clinical recurrence, and investigated whether ctDNA clearance following adjuvant therapy reflects treatment efficacy.

## Methods

### Ethics statement

All patients provided written informed consent. The protocol was approved by the Dana Farber/Harvard Cancer Center institutional review board, registered on ClinicalTrials.gov (NCT06730412) and was conducted in accordance with the Declaration of Helsinki and in compliance with the U.S. Common Rule.

### Study design and participants

We conducted a prospective observational cohort study, Clear-HPVca (ctDNA Liquid Biopsy for Early Assessment of Residual Disease in HPV-associated Head and Neck Cancer) that enrolled 103 AJCC 8 Stage I-III HPV+HNC patients at Massachusetts Eye and Ear/Massachusetts General Hospital between August 2020 and March 2024. Key eligibility criteria were: 1) age ≥18 years, 2) newly diagnosed, untreated, histologically confirmed HPV+HNC of any head and neck anatomic site, 3) scheduled for curative intent resection as primary treatment, 4) Eastern Cooperative Oncology Group (ECOG) performance status of 0-2. HPV status was confirmed by histomorphology consistent with squamous cell carcinoma on tissue biopsy and p16 immunohistochemistry with or without HPV PCR, or HPV RNA in situ hybridization per College of American Pathology guidelines^30^. Adjuvant therapy after surgery was determined by a multidisciplinary tumor board based on clinicopathologic risk factors. Clinical post-treatment examinations occurred at 2-to 4-month intervals per National Comprehensive Cancer Network (NCCN) guidelines and included physical exam and endoscopy. Cross-sectional imaging was obtained at three months and one year post-treatment completion, as well as at the additional discretion of the treating physician. Clinical information was extracted from the Electronic Health Record and input into a study-specific electronic data capture system. Three independent reviewers extracted data from surgical pathology reports to ensure accuracy, and conflicts were manually re-reviewed to ensure accuracy and consistency.

Three MRD windows were analyzed: 1) MRD immediately post-surgery (MRD-early) was defined as post-operative days 1-3, 2) MRD post-surgery (MRD-PS) was defined as 4 days to 6 months after surgery and before the start of adjuvant therapy, 3) MRD post-treatment completion (MRD-TC) was defined as 4 days to 6 months after surgery for patients treated with surgery alone and <6 months after radiation completion for patients treated with adjuvant radiation with or without chemotherapy. The potential clinical application of analysis in these windows includes: 1) for MRD-early, immediate prognostication and personalized adjuvant treatment selection, 2) for MRD-PS, presumed optimal prognostication and personalized adjuvant treatment selection timing, and 3) for MRD-TC prognostication and personalized post-adjuvant treatment and surveillance. Post-operative day (POD) 4 was chosen as the earliest time point following surgery for the MRD-PS window as a separate analysis was conducted examining POD 1-3 vs POD 4-14 values in patients treated with surgery in a lead-in cohort demonstrating clearance occurred in some patients at the later time points with this more sensitive assay, contrary to earlier data our group had generated examining POD 1 as a landmark timepoint with ddPCR-based analysis^23, 31^. The surveillance window was defined as the time from the end of the MRD-TC window to the last follow-up, recurrence, or death. The relevant clinical application is the early detection of recurrence, which could trigger non-routine imaging and/or salvage treatment initiation.

### ctHPVDNA testing

A clinically validated multi-feature HPV whole genome sequencing assay, HPV-DeepSeek, was used for quantification of ctHPVDNA. HPV-DeepSeek uses hybrid capture NGS to detect and quantify 43 HPV genotypes, mutations in PIK3CA, viral integration events, prognostic viral single-nucleotide polymorphisms (SNPs), and multiple fragment size features. Detailed HPV-DeepSeek methodology can be found in Bryan et. al^8^. Samples were considered positive using a pre-defined threshold (≥10 unique HPV reads and ≥10% HPV genome coverage), as described previously^8^. A clinically validated ddPCR assay targeting specific regions of the E7 gene from five high-risk HPV genotypes: 16, 18, 33, 35, and 45 was used for head-to-head comparisons to HPV-DeepSeek^8, 19, 23, 24, 31–34^. Samples were considered positive using a pre-defined threshold (≥ 2 HPV reads), as described previously^9^.

## Statistical Analysis

The primary endpoints were disease-free survival (DFS) and overall survival (OS). The primary hypothesis tested was that patients with MRD after surgery (MRD-PS window) would have inferior DFS and OS compared to patients without MRD. The secondary endpoint was DFS and OS after treatment completion (MRD-TC window) in patients with and without MRD. DFS was defined as the period from the date of treatment completion (surgery or the completion of adjuvant therapy) to the date of relapse. OS was defined as the time from treatment completion to the date of death from any cause. The Kaplan–Meier (KM) method was used to estimate the survival distribution. Differences between groups were tested using the log-rank test. When multiple samples were taken within a designated window, any positive sample was regarded as MRD positive. Disease recurrence required confirmatory tissue biopsy.

The exploratory endpoints included: (1) DFS and OS in patients with and without MRD in the MRD-early window, (2) timing of detection of molecular recurrence compared to clinical detection of recurrence, (3) sensitivity and lead times of HPV-DeepSeek compared to ddPCR, and (4) identification of key features driving recurrence risk. Prognostic factors associated with DFS were evaluated using a multivariable Cox proportional hazard model.

To assess the ability of clinical and molecular features to predict recurrence over time, we fit univariate Cox proportional hazards models using time from biopsy to recurrence or last follow-up as the outcome. Patients without recurrence were right-censored at their last clinical follow-up. Predictors included ctHPVDNA status during the MRD-PS window, extranodal extension (ENE), surgical margin status, and involvement of more than two lymph nodes. Model discrimination was assessed using Harrell’s concordance index (C-statistic).

To compare predictive accuracy between variables, we calculated differences in C-statistics (ΔC) with 1,000 bootstrap samples to estimate standard errors and 95% CIs. KM curves were generated for visual comparison of DFS across groups, with log-rank tests used to assess statistical significance. Analyses were performed in R (v4.4.1) in a Jupyter Notebook.

## Results

### Patient cohort and characteristics

103 patients were enrolled over a 44-month period (8/18/2020-3/14/2024) (Figure 1). A total of 560 samples were collected and processed (mean of 5.6 samples/patient). The mean age was 62 years (range 36-84). The cohort was predominately male (89%: 92/103), AJCC8 stage I (89%: 92/103) and oropharyngeal subsite (93%: 96/103) (Table 1, Supplementary Table 1). HPV16 was the dominant genotype detect by HPV-DeepSeek (88%: 89/101), with 5 additional genotypes detected (Figure 2A).

**Figure 1.**
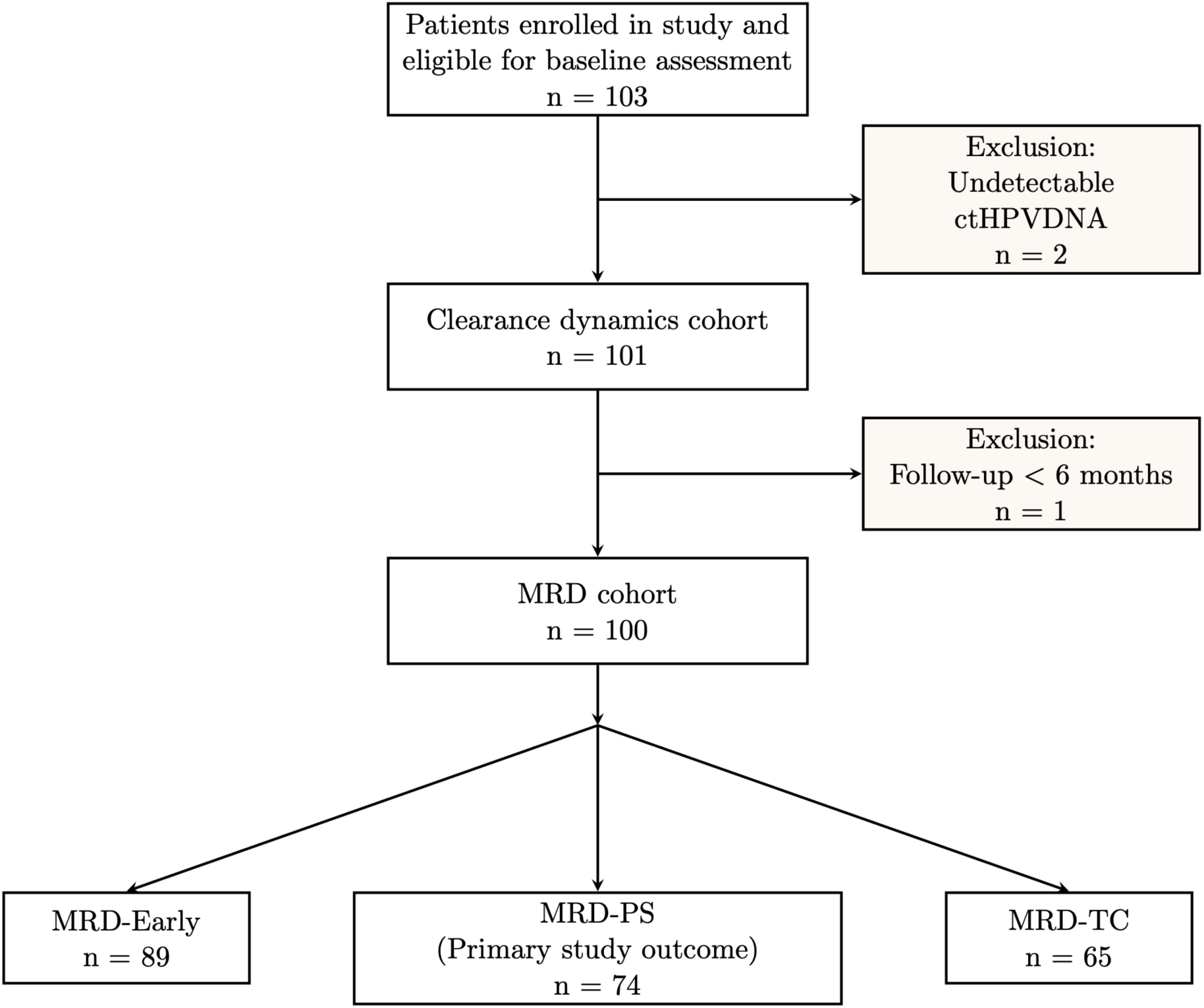
CONSORT diagram of study population selection and outcome groups. The diagram outlines progression from the total enrolled population through exclusions to the clearance dynamics cohort, the MRD analysis cohort, and the three defined MRD windows: MRD-Early, MRD-PS (primary study outcome), and MRD-TC.

**Figure 2.**
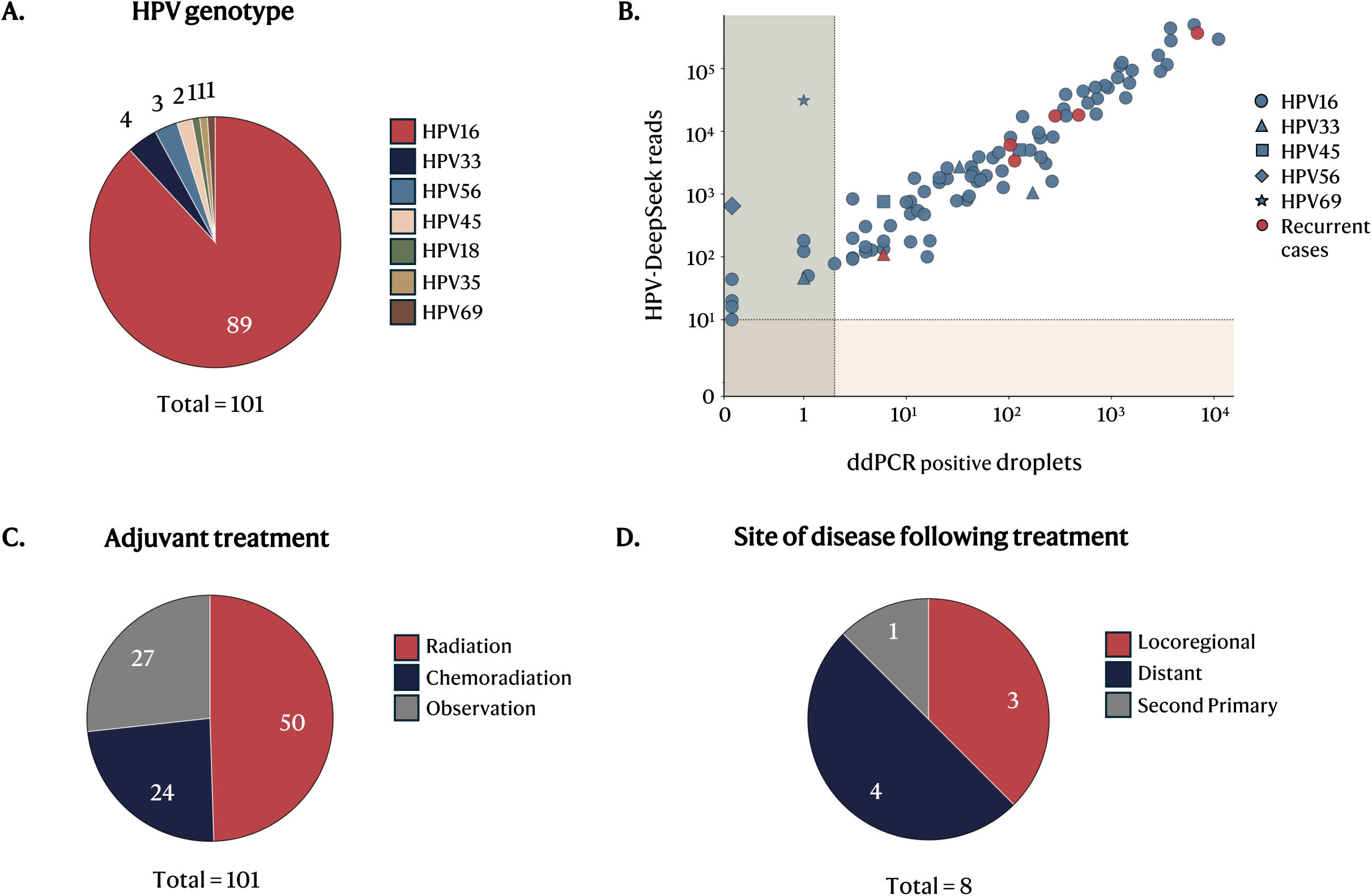
Patient characteristics and ctHPVDNA detection at diagnosis (**A**) HPV genotype detected by HPV-DeepSeek. (**B**) Correlation between HPV-DeepSeek (y-axis) and ddPCR (x-axis) in 92 cases run on both assays prior to treatment. Red dots represent cases with recurrence. The shape of each dot represents the HPV genotype. The shaded area represents the HPV-DeepSeek (yellow) and ddPCR (green) negative cases. (**C**) Adjuvant treatment across the cohort. (**D)** Site of disease following treatment.

**Table 1.** Patient demographics, staging, and clinicopathologic risk factors. (A) Clinical staging and demographic characteristics of 103 HPV+HNC cases treated with surgery. **(B)** Pathological staging and clinicopathologic risk factors of 101 cases, which were ctHPVDNA positive at the baseline and included in the survival analysis.

| Pre-operative Characteristic | Patients No. (%)<br>n = 103 |
| --- | --- |
| Age at diagnosis, mean (range) | 62 (36–84) |
| Sex |  |
| Male | 92 (89) |
| Female | 11 (11) |
| Smoking status |  |
| Never smoked | 49 (48) |
| <10 pack/year | 21 (20) |
| >10 pack/year | 33 (32) |
| Clinical AJCC 8 stage |  |
| I | 92 (89) |
| II | 10 (10) |
| III | 0 (0) |
| IV | 1 (1) |
| Clinical T stage |  |
| 0 | 18 (17) |
| 1 | 49 (48) |
| 2 | 32 (31) |
| 3 | 3 (3) |
| 4 | 1 (1) |
| Clinical N stage |  |
| 0 | 17 (17) |
| 1 | 82 (80) |
| 2 | 4 (4) |
| 3 | 0 |
| Clinical M stage |  |
| 0 | 103 (100) |
| 1 | 0 |
| Viral genotype at diagnosis |  |
| HPV16 | 14 (14) |
| HPV16/18 | 74 (72) |
| Other high risk | 11 (11) |
| Unknown | 4 (4) |
| Primary site |  |
| Tonsil | 48 (47) |
| Base of tongue | 48 (47) |
| Oral cavity | 2 (2) |
| Nasal cavity | 2 (2) |
| Unknown primary | 2 (2) |
| Overlapping | 1 (1) |

**B.**
| Post-operative Characteristic | Patients No. (%)<br>n = 101 |
| --- | --- |
| AJCC 8 pathological stage |  |
| I | 89 (88) |
| II | 10 (10) |
| III | 0 |
| IV | 2 (2) |
| Pathological T stage |  |
| 0 | 2 (2) |
| 1 | 47 (47) |
| 2 | 47 (47) |
| 3 | 3 (3) |
| 4 | 2 (2) |
| Pathological N stage |  |
| 0 | 13 (13) |
| 1 | 81 (80) |
| 2 | 3 (3) |
| 3 | 0 |
| Unknown | 4 (4) |
| Number of lymph nodes |  |
| 0–1 | 61 (60) |
| ≥2 | 36 (36) |
| Unknown | 4 (4) |
| Extranodal extension |  |
| Positive | 28 (28) |
| Negative | 70 (69) |
| Unknown | 3 (3) |
| Margin status |  |
| Positive | 7 (7) |
| Negative | 90 (89) |
| Unclear / Unknown | 4 (4) |
| Perineural invasion |  |
| Positive | 8 (8) |
| Negative | 91 (90) |
| Unknown | 2 (2) |
| Lymphovascular invasion |  |
| Positive | 29 (29) |
| Negative | 69 (68) |
| Unknown | 3 (3) |

### ctHPVDNA detection with HPV-DeepSeek at clinical diagnosis

The mean pre-operative ctHPVDNA level was 32,617 (range 8-495,758), with a mean HPV genome coverage of 85.3% (range 8.5%-100%). The sensitivity of HPV-DeepSeek was 98.1% (101/103). 92 cases had both HPV-DeepSeek and ddPCR performed with sensitivities of 99% (91/92) and 89% (82/92), respectively. There was a linear correlation between ddPCR and HPV-DeepSeek reads (Pearson correlation coefficient (r)=0.86, P<0.001) (Supplementary Figure 1). HPV-DeepSeek positive, ddPCR negative samples had low ctHPVDNA levels or genotypes outside the ddPCR panel, leading to significantly improved sensitivity in HPV-DeepSeek vs ddPCR (p=0.004) (Figure 2B).

### MRD cohort and characteristics

HPV-DeepSeek negative cases at enrollment were excluded from subsequent analysis (n=2). 73% (74/101) of patients received adjuvant therapy after surgery: 50% (50/101) received radiation therapy, and 24% (24/101) received chemoradiotherapy (Figure 2C). Seven patients experienced a recurrence (4 distant and 3 locoregional), and one patient was found to have a second HPV+ primary (sinonasal) cancer during the surveillance period (Figure 2D). There were six deaths. The median follow-up was 796 days (range: 138-1376 days).

### ctHPVDNA clearance dynamics after treatment

We first evaluated how baseline ctHPVDNA levels impacted ctHPVDNA clearance. For this analysis patients were grouped into four categories: (1) those who became MRD negative in the MRD-early window, (2) those who became MRD negative in the MRD-PS window, (3) those who became MRD negative after adjuvant treatment, and (4) those who remained MRD positive following adjuvant treatment (Figure 3A). The rapid clearance group (cleared by the MRD-early window) had significantly lower baseline ctHPVDNA values than those who cleared more gradually (cleared by MRD-PS) (p=0.0037) and those who cleared only after adjuvant treatment (MRD-TC) (p=0.0001).

**Figure 3.**
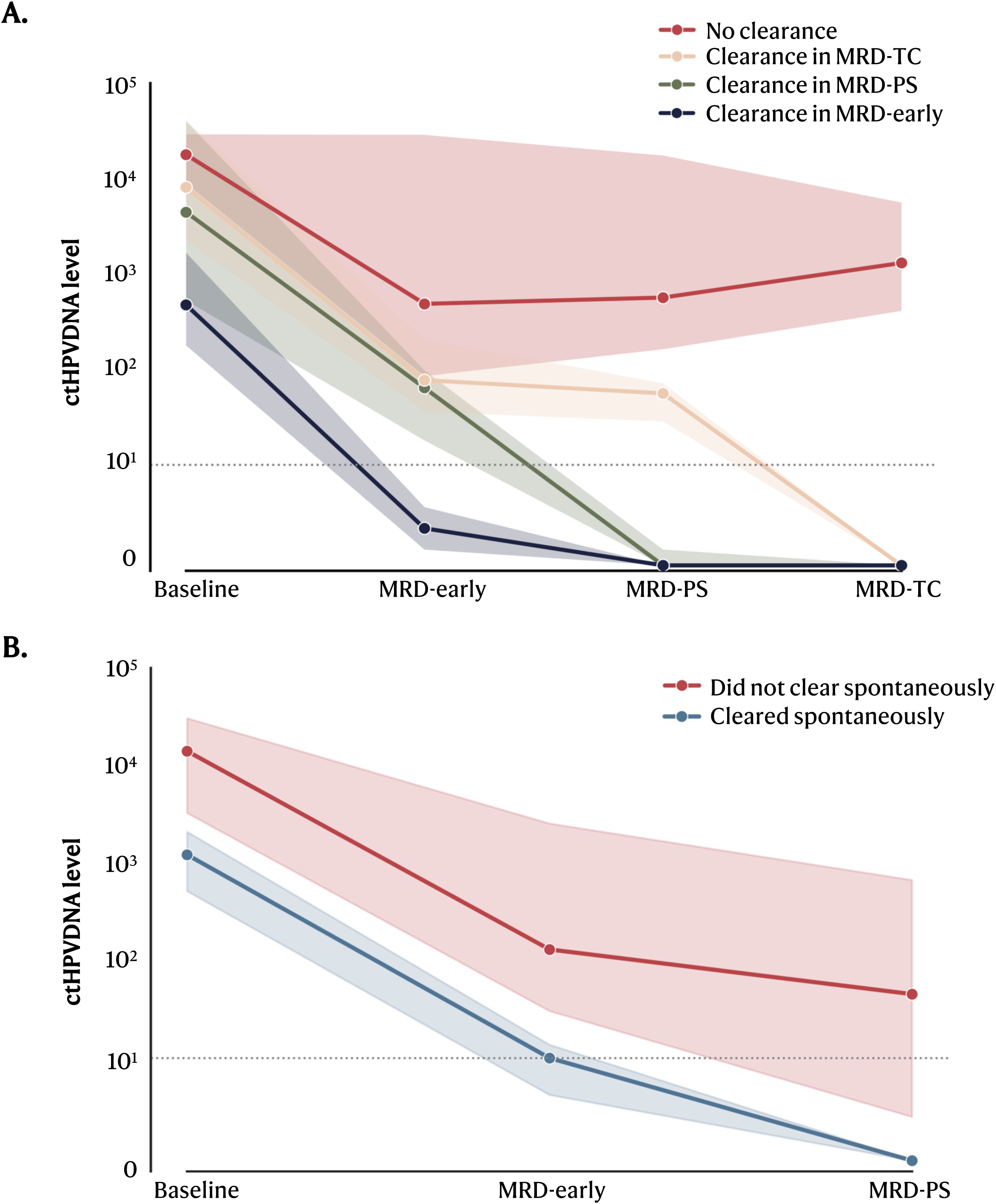
ctHPVDNA clearance dynamics. **(A)**. Dynamics of ctHPVDNA after surgery. Cases were divided into four groups based on timing of ctHPVDNA clearance: 1. No clearance (red), 2. Clearance in MRD-TC (clearance after adjuvant treatment) (yellow), 3. Clearance in MRD-PS (Gradual clearance) (green), 4. Clearance in MRD-Early (rapid clearance) (blue). The line and shades represent the median ctHPVDNA level and interquartile range, respectively. **(B).** ctHPVDNA decay from baseline to MRD-early and MRD-early to MRD-PS. Patients were grouped into two categories: (1) cleared spontaneously (cleared by MRD-early or MRD-PS, i.e. before adjuvant treatment) (red line) or did not clear spontaneously (ie, cleared after adjuvant therapy or remained positive) (teal line).

To further characterize clearance dynamics, we modelled per-patient ctHPVDNA decay from baseline to MRD-early and MRD-early to MRD-PS. For this analysis, patients were grouped into two categories: (1) cleared spontaneously (cleared by MRD-early or MRD-PS, i.e., before adjuvant treatment) or did not clear spontaneously (i.e., cleared after adjuvant therapy or remained positive). While the clearance slope from baseline to MRD-early was similar between groups, the clearance slope from MRD-early to MRD-PS diverged between groups. During this interval, patients who cleared spontaneously had a significantly steeper median slope (–0.060 vs 0.0049, FDR-adjusted p=0.040), suggesting that slower clearance in this time window could be an additional metric used to predict MRD status in addition to ctHPVDNA value (Figure 3B, Supplementary Table 2).

### Association of ctHPVDNA status in MRD windows with recurrence and survival

One patient was excluded from the recurrence and survival analysis due to follow-up <6 months, leaving 100 patients for analysis. ctHPVDNA status was available for 74 patients during the MRD-PS window, the primary outcome assessed. 17/74 patients (23%) were MRD positive, with six having a recurrence. 57/74 patients (77%) were MRD negative with one having a recurrence. The single recurrence in the MRD negative group occurred at 31 months, leading to a 2-year DFS of 60% (95% CI: 31-80) vs 100% respectively (p<0.001) (Figure 4A). The association of MRD positivity with a significantly increased risk for recurrence was observed for both the earliest stages of disease (T0-1 N0-1M0 2-year DFS 80% (95% CI: 20-97) vs 100%, p=0.046) and later stages (55% (95% CI: 23-78) vs 100%, p<0.001) as well as when restricted to only oropharynx cancer (55% (95% CI: 26-77) vs 100%, p<0.001) (Supplementary Figures 2A-C). MRD positivity was also found to be significantly associated with worse OS compared to MRD negative patients, demonstrating a 2-year OS of 73% (95% CI: 43-89) vs 98% (95% CI: 88-100), respectively (p<0.002) (Figure 4B). The association of MRD positivity with increased risk of death was observed for the later stages (2-year OS 60% (95%CI: 24-83) vs 100%, p=0.001) as well as when restricted to only oropharynx cancer (70% (95%CI: 38-88) vs 98% (95%CI: 88-100), p=0.001) (Supplementary Figures 3 A-B). There was no association with OS at the earliest stages of disease (T0-1N0-1M0 2-year OS 100% vs 98% (95% CI:85-100), p=0.65). Restricting the analysis to only the earliest timepoint in the window, when multiple timepoints were available, showed slightly lower HRs, suggesting that multi-timepoint sampling within a window increases diagnostic accuracy for MRD detection, but results were largely unchanged (Supplementary Table 3, Supplementary Figure 4 A-D).

**Figure 4.**
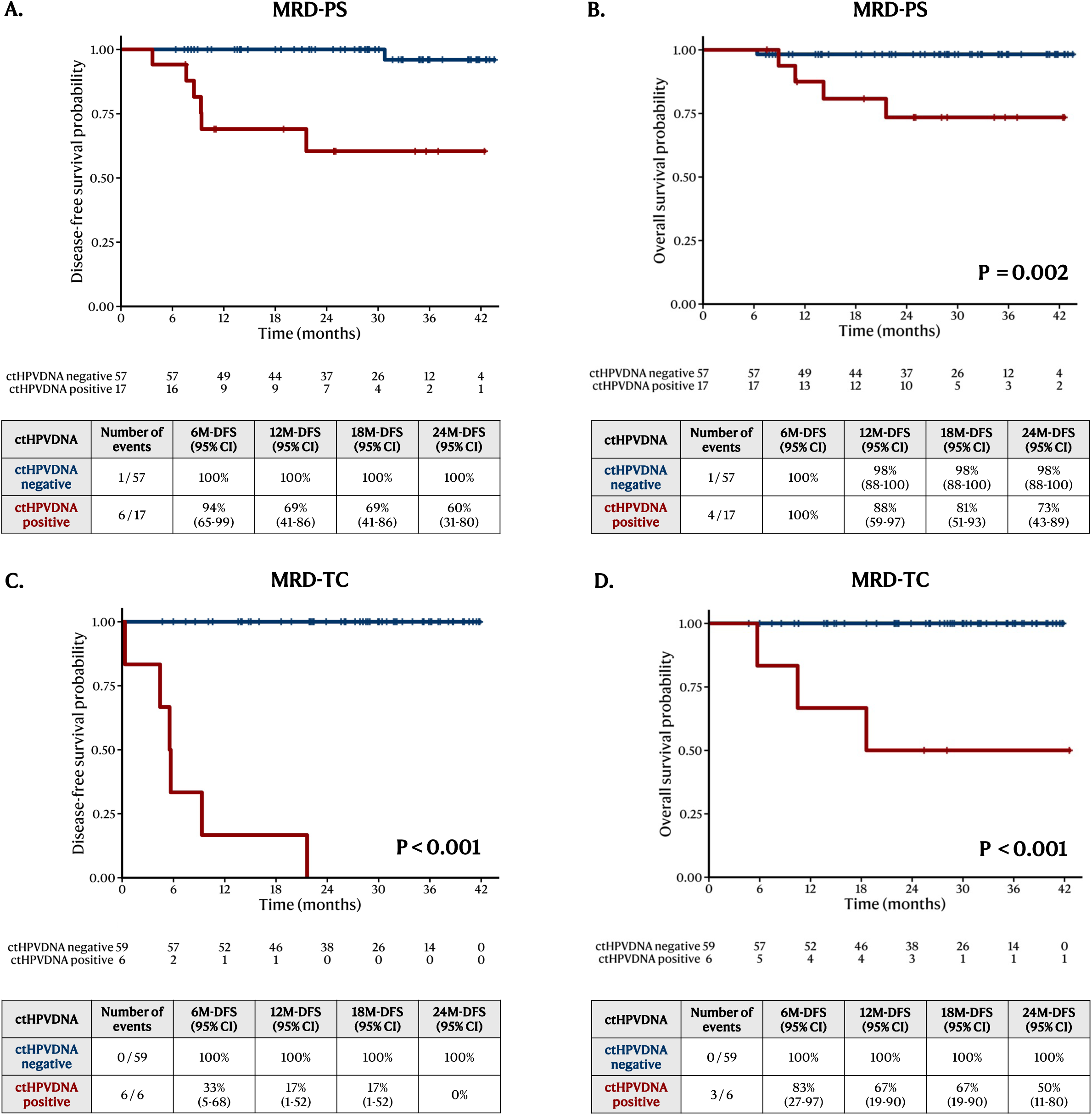
Association of ctHPVDNA with recurrence and survival (**A, B**) Kaplan-Meier curve of two-year DFS and OS based on ctHPVDNA status during the MRD-PS window. The X-axis shows months after surgery, and the Y-axis shows the recurrence/survival rate. The red line represents ctHPVDNA positive, and the blue line represents ctHPVDNA negative. The Log-rank test was used to calculate the P value. The number at risk every 6 months is displayed at the bottom of the graph. (**C, D**) DFS and OS based on MRD status in the MRD-TC window.

ctHPVDNA status was available for 65 patients during the MRD-TC window, the secondary outcome assessment. 6/65 (9%) were MRD positive, with six (100%) having a recurrence. 59/65 (91%) were MRD negative, with 0% (0/59) having a recurrence, demonstrating a 2-year DFS of 0% vs 100% respectively (p<0.001) (Figure 4C). The association of MRD positivity with a significantly increased risk for recurrence was observed for both the earliest stages of disease (T0-1N0-1M0 2-year DFS (0% vs 100%, p<0.001) and later stages (0% vs 100%, P<0.001) as well as when restricted to only oropharynx cancer (DFS 0% vs 100%, p<0.001) (Supplementary Figure 5A-C). MRD positivity was also found to be significantly associated with worse OS compared to MRD negative patients, demonstrating a 2-year OS of 50% (95% CI: 11-80) vs 100% respectively (p<0.001) (Figure 4D). The association of MRD positivity with a significantly increased risk for death was observed for the later stages (2-year OS 40% (95%CI: 5-75) vs 100%, p<0.001) as well as when restricted to only oropharynx cancer (50% (95%CI:11-80) vs 100%, p<0.001) (Supplementary Figure 6A-B). In the earliest stages of disease (T0-1N0-1M0), there were no deaths regardless of MRD status.

ctHPVDNA status was available for 89 patients during the MRD-early window, an exploratory outcome. 48/89 (54%) were MRD positive with seven having a recurrence. 41/89 (46%) were MRD negative with one having a recurrence, demonstrating a 2-year DFS of 87% (95% CI: 73-94) vs 96% (95% CI: 76-99) respectively (p=0.052) (Supplementary Figure 7A). MRD positivity was also not associated with worse OS compared to MRD negative patients demonstrating a 2-year OS of 88% (95% CI: 74-95) vs 97% (95% CI: 81-100) respectively (p=0.18) (Supplementary Figure 7B).

### HPV-DeepSeek versus ddPCR for detecting MRD, recurrence and determining type of recurrence

We compared the sensitivity and lead times from molecular detection of recurrence to clinical detection of recurrence using HPV-DeepSeek and currently clinically available HPV liquid biopsy (ddPCR). To evaluate sensitivity, we examined patients with biopsy confirmed recurrence (7) and utilized MRD status in the MRD-PS window. HPV-DeepSeek sensitivity was 83% (5/6) vs 33% (2/6) with ddPCR (Figure 5A), and when restricting to only the first timepoint, if multiple timepoints were sampled in the window, sensitivity was 67% (4/6) vs 33% (2/6), respectively. In the MRD-TC window, HPV-DeepSeek sensitivity was 100% (5/5) vs 60% (3/5) with ddPCR (Figure 5B), and when restricting to only the first time point, sensitivity was 60% (3/5) vs 40% (2/5), respectively.

**Figure 5.**
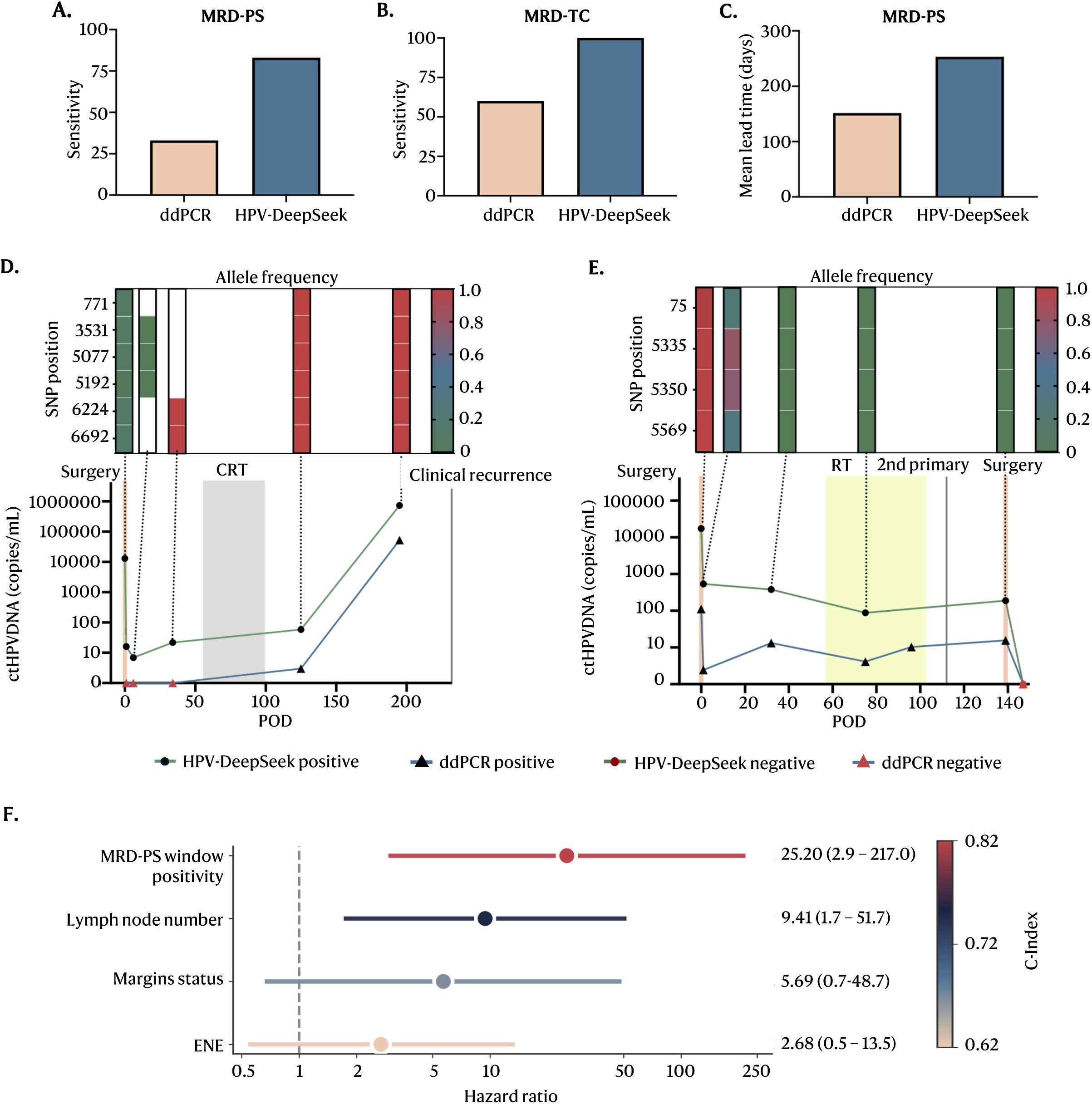
HPV-DeepSeek versus ddPCR for detecting MRD, recurrence, and determining type of recurrence. (**A, B**) Bar graph showing the sensitivity of HPV-DeepSeek and ddPCR for MRD after surgery (MRD-PS) and treatment completion (MRD-TC). The y-axis shows sensitivity (%). (**C**) Bar graph showing the mean lead time to clinical diagnosis of recurrence after surgery by HPV-DeepSeek and ddPCR assay. Line graphs showing the changes in ctHPVDNA levels over time for a recurrent case (**D**) and 2^nd^ primary case (**E**). Each plot represents one patient’s ctHPVDNA trajectory as a function of time, with the sample collection timepoint in days after surgery on the x-axis and the ctHPVDNA levels on the y-axis (logarithmic scale). The green line represents the ctHPVDNA value transformed to copies/ml for HPV-DeepSeek, and the blue line for the ddPCR. Black circles are positive samples and red triangles are negative samples. The allele frequency (AF) heatmap of viral SNPs is shown above graph with green and red representing an AF of 0 and 1 at each SNP position, respectively. The vertical axis shows the genome position of each SNP, and the horizontal axis shows the time course. The vertical dotted lines connect each sample (bottom) with the AF (top). The timing of surgery is shown with red vertical line. The timing of chemoradiation is shown with a grey shaded box. The timing of radiation is shown with a yellow box. **(F).** Forest plot showing hazard ratios (HR) and 95% confidence intervals for disease-free survival by prognostic factor, with color scale indicating Harrell’s concordance index (C-index) for each variable. MRD-PS window positivity, lymph node number, margin status, and extranodal extension (ENE) are displayed, with HR plotted on a logarithmic scale.

The mean time to biopsy-proven recurrence in the cohort was 447 days. To measure lead time, we utilized the first positive sample beginning in the MRD-PS window. The mean lead time was 214 days (median 155, range 59-533) by HPV-DeekSeek and 125 days (median 83, range 0-421) by ddPCR (Figure 5C, Supplementary Table 4).

To investigate if recurrence type could be differentiated, we performed viral genome fingerprinting across all patients with longitudinal positive samples after treatment completion (n=7). Three patients had locoregional recurrences, three patients had distant recurrences, and one patient had a second primary HPV+HNC (sinonasal) (Supplementary Figure 8A-G). Five patients had identical viral genome fingerprints across all samples within a patient, while two patients had the emergence of new viral genome fingerprints. The first patient had a six SNP fingerprint present at diagnosis which was replaced by a new fingerprint by POD 125. An admixture of these fingerprints was seen in the post-operative windows (Figure 5D). This patient was clinically diagnosed with liver metastases on POD 232, suggesting the fingerprint evolution represented the extirpation of the tumor with surgery and the emergence of the metastatic lesion, differentiated by the unique viral SNPs arising from a viral subclone. In a second patient, a four-SNP fingerprint was present at diagnosis, which was replaced by a new fingerprint by POD 32. An admixture of these fingerprints was seen one day after surgery (Figure 5E). A second primary cancer was diagnosed in the sinonasal cavity on POD 112, suggesting the fingerprint evolution represented the new, unique viral genome. This finding was validated using tumor tissue sequencing from both the first and second primary cancers.

### MRD status as an independent predictor of recurrence and benefit from adjuvant therapy

MRD-PS window positivity was the strongest univariate predictor of DFS (HR 25.2; 95% CI: 2.93–217.0; p=0.003) and had the highest discriminative performance (C=0.822) compared to top the traditional clinicopathologic risk factors such as number of lymph nodes (HR 9.41; 95% CI: 1.71–51.7; p=0.010; C=0.751), margin status (HR 5.69; 95% CI: 0.66–48.7; p=0.112, C=0.669) and presence of ENE (HR 2.68; 95% CI: 0.54–13.45; p=0.230; C=0.622) (Figure 5F)^35^.

We examined the impact of adjuvant treatment on both MRD positive and MRD negative patients in the MRD-PS window, adjusting for stage. 23% (17/75) of patients were MRD positive, of whom 88% (15/17) received adjuvant treatment and 77% (58/75) of patients were MRD negative, of whom 64% (37/58) received adjuvant treatment. MRD positive patients benefited from adjuvant treatment with a recurrence rate of 27% (4/15) for the adjuvant therapy group versus 100% (2/2) for the no-adjuvant therapy group (adjusted HR: 0.162, 95% CI: 0.026-1.009, P=0.05). No benefit was observed for MRD negative patients with a recurrence rate of 3% (1/37) in the adjuvant therapy group versus 0% (0/21) for the no-adjuvant therapy group) (P=0.997).

We next assessed if ctHPVDNA clearance following adjuvant treatment (MRD-TC and Surveillance windows) was predictive of the efficacy of adjuvant treatment, as well as outcomes. Of the 17 patients who were MRD positive in the MRD-PS window, patients who cleared ctHPVDNA after adjuvant treatment had superior DFS and OS (DFS: 100% vs 0%, P<0.001; OS: 100% vs 50% (95%CI: 11.1-80.4), P=0.039).

## Discussion

Accurate risk stratification following surgery in HPV+HNC is a major unmet clinical need driven by an increasing disease incidence and a common treatment strategy of surgery followed by risk-adjusted adjuvant treatment. Currently, decisions regarding adjuvant therapy rely on clinicopathologic features, which perform poorly in identifying MRD on an individual basis. Studies evaluating the prognostic significance of ctHPVDNA after surgery, which is in use nationally as a surveillance tool for HPV+HNC following treatment completion, have found suboptimal performance using existing ddPCR-based assays due to limited sensitivity^23–27^. In this prospective study of 103 HPV+HNC patients treated with curative-intent surgery and followed for a median of 27 months, we demonstrate that the application of an ultrasensitive HPV WGS assay enables accurate MRD detection with strong prognostic and predictive implications. Principally, we show that the presence of ctHPVDNA both after surgery and following completion of adjuvant therapy is associated with a markedly increased risk of recurrence and death. HPV WGS outperformed ddPCR in both sensitivity and lead time to recurrence detection and emerged as the most powerful prognostic factor associated with both DFS and OS compared to currently utilized clinicopathologic features. These findings support the integration of HPV WGS ctHPVDNA analysis into future prospective interventional trials aimed at refining postoperative risk stratification and guiding treatment personalization for HPV+HNC patients treated with surgery.

Our results build on prior work demonstrating the potential utility of MRD detection in the postoperative setting in HPV+HNC. In the first study to assess the role of ctHPVDNA in MRD detection O’Boyle et al. demonstrated that ctHPVDNA-detection by ddPCR cleared rapidly (within 24 hours) following surgery in patients with complete tumor extirpation but remained elevated in patients with macroscopic residual disease^23^. However, in this work, and subsequent ddPCR-based studies, ctHPVDNA demonstrated limited sensitivity in the context of microscopic residual disease^24–27^. In the secondary analysis of the phase III DART trial, Routman et al. found that postoperative ctHPVDNA was associated with poorer DFS and OS, but some ctHPVDNA-negative patients with advanced nodal disease experienced high recurrence rates^25^. Similarly, Chen et al. reported that in a prospective trial, 25% of ctHPVDNA-negative patients experienced early recurrence, leading to closure of the trial, highlighting the insufficiency of existing methods to reliably guide adjuvant treatment decisions.

Our study directly addresses these limitations by prospectively validating a novel HPV WGS assay, HPV-DeepSeek, which was previously shown to have markedly improved sensitivity over ddPCR when applied at the time of cancer diagnosis^8^. These findings echo emerging human WGS-based assays which show improved sensitivity over more targeted panel or WES-based ctDNA assays, but with the advantage of HPV WGS assays having significantly reduced turnaround time and cost compared to human WGS^8, 36, 37^.

We first evaluated HPV-DeepSeek performance at trial enrollment. Current studies evaluating ddPCR-based MRD detection have demonstrated that 14-40% of patients have low or negative ctHPVDNA at trial enrollment, precluding accrual to the trial completely.

Here, HPV-DeepSeek demonstrated high sensitivity at the time of cancer diagnosis, as shown previously, with <2% false negatives at trial enrollment, permitting >98% of patients to enroll.

To assess MRD capacity, we first evaluated optimal MRD sampling, using multiple postoperative windows for MRD assessment as well as single vs multi-timepoint sampling within the windows. While our group had previously shown that early postoperative sampling is possible using ddPCR (POD 1), here we found sampling within the first 3 days may confound results when using a significantly more sensitive approach such as HPV WGS^23^.

This timing aligns well with standard postoperative surveillance visits. We further found repeated sampling (multi-timepoint sampling) increased the sensitivity of MRD assessment, suggesting that future prospective trials should include multiple MRD timepoints within the MRD window to optimize diagnostic accuracy. We also examined ctHPVDNA clearance dynamics finding that that initial ctHPVDNA values influence the time to clearance, and separately, that the slope of clearance is associated with the likelihood of residual disease and thus the likelihood of requiring adjuvant therapy.

Our key findings show that patients with detectable MRD after surgery are at markedly higher risk of recurrence and death. In contrast, patients without detectable MRD had near-universal favorable outcomes regardless of whether they received adjuvant therapy.

Importantly, ctHPVDNA status also predicted benefit from adjuvant treatment: patients who cleared MRD after therapy had excellent outcomes, while those with persistent ctHPVDNA all recurred. This strongly supports a framework in which MRD status not only informs prognosis but also acts as a predictive biomarker for treatment response. This mirrors findings in colorectal cancer and lung cancer MRD studies, where ctDNA clearance after therapy strongly predicts cure^11, 38^.

In our study, ctHPVDNA-based MRD status outperformed conventional features in predicting recurrence, supporting its potential use alone or in combination with existing indications to personalize adjuvant therapy. For treatment escalation, MRD positivity identified patients at high risk of recurrence even in the absence of standard high-risk pathologic findings, supporting a rationale for adjuvant therapy in this setting. Conversely, for treatment de-escalation, patients with standard indications for chemoradiotherapy (e.g., ENE) but no MRD had a very low recurrence risk, suggesting that less intensive therapy, for example, radiation alone, may be sufficient. Prospective trials will be essential to validate these strategies.

These concepts are further supported by our finding that MRD-negative patients derived no survival benefit from adjuvant therapy, whereas MRD-positive patients had improved outcomes with adjuvant treatment, establishing ctHPVDNA MRD status as a predictive biomarker of therapeutic benefit.

Ongoing clinical trials are exploring ctHPVDNA-guided de-escalation strategies in HPV-associated head and neck cancer, primarily using ddPCR-based assays. In the Winship-5566 trial, adjuvant radiotherapy dose is tailored based on MRD status: patients who are MRD-negative receive 36 Gy, while MRD-positive patients with intermediate-risk features receive 50–60 Gy. Similarly, in the DART 2.0 trial, MRD-negative patients with a single intermediate-risk factor are observed without adjuvant therapy, while those with pN1 disease may be de-escalated if MRD-negative. Notably, in our study, HPV-DeepSeek identified MRD in a subset of patients who were negative by ddPCR, demonstrating nearly double the sensitivity. While these trials represent important steps toward treatment personalization, our findings highlight that more sensitive assays—such as those used here—may be required to safely enable de-escalation strategies without compromising oncologic outcomes.

The implications of more sensitive ctHPVDNA detection extend beyond guiding adjuvant therapy. In a recent prospective surveillance study in HPV+HNC, Rettig et al. reported that ddPCR-based ctHPVDNA testing detected only 40% of recurrences prior to standard-of-care imaging or physical examination, with an average lead time of only ∼2 months^39^. In contrast, HPV-DeepSeek demonstrated a substantially longer interval between molecular and clinical recurrence—averaging over seven months and extending up to 17.5 months—nearly double the four-month lead time observed with ddPCR in our cohort. This extended window for early detection creates new opportunities for preemptive intervention and may improve salvage outcomes. Moreover, ctHPVDNA WGS analysis has the potential to resolve diagnostic challenges in the surveillance setting, such as distinguishing recurrence from a second primary tumor, as shown by viral fingerprinting in this cohort.

The ability to directly assess MRD in real time offers a potential paradigm shift: moving from static, probabilistic estimates to dynamic, patient-specific risk assessment. This opens the door for prospective trials testing MRD-guided treatment de-escalation or intensification—ultimately reducing unnecessary toxicity while preserving oncologic outcomes. Similar principles have been successfully applied in other malignancies. In colorectal cancer, for instance, Tie et al. showed that MRD detection via ctDNA post-resection was associated with recurrence risk and could inform adjuvant chemotherapy decisions^40^. In lung cancer, the DYNAMIC study and other MRD-focused trials are now prospectively evaluating ctDNA-guided adjuvant therapy. Our findings suggest that HPV+HNC is equally suited for this approach, and future trials should explicitly evaluate MRD-guided adjuvant treatment using HPV WGS approaches.

While our cohort was prospectively enrolled and rigorously followed, the recurrence event rate remains low, reflecting the generally favorable prognosis of HPV+HNC. Nonetheless, the strength and consistency of the associations observed lend confidence to the findings. Further, the study was conducted at a single high-volume academic center, and external validation in multicenter trials, including community sites, and in larger and more diverse populations is warranted to confirm generalizability and inform clinical implementation. In this trial, we included all HPV+HNC, not only oropharynx cancers, increasing the generalizability. However, while we could validate the prognostic role of MRD in subgroup analysis of oropharynx cancers alone, the small sample size of non-oropharynx cancers precluded statistical power in these other sites. Importantly, we have previously shown ctHPVDNA detection appears to perform equally in non-oropharynx subsites^31^. Similarly, most cases were early stage, precluding subgroup analysis of advanced stage cancers. Importantly, surgery for advanced stage HPV+HNC is rare.

In conclusion, this study provides strong clinical evidence that ctHPVDNA MRD—when assessed using an ultrasensitive WGS assay—predicts recurrence and survival following surgery for HPV+HNC and may inform adjuvant therapy decisions. HPV-DeepSeek enabled accurate risk stratification, early detection of recurrence, and prediction of therapeutic response, representing an advance toward precision oncology in HPV+HNC. ctHPVDNA-based MRD detection has the potential to transform the management of HPV+HNC by enabling precise, personalized, and dynamic risk stratification. These findings support the potential for a shift from static pathologic criteria toward a more nuanced, biology-driven model—one that aligns treatment intensity with individual disease risk and thereby improves both survival and quality of life.

## Data Availability

All data produced in the present work are contained in the manuscript

---

**Supplementary Figure 1.**
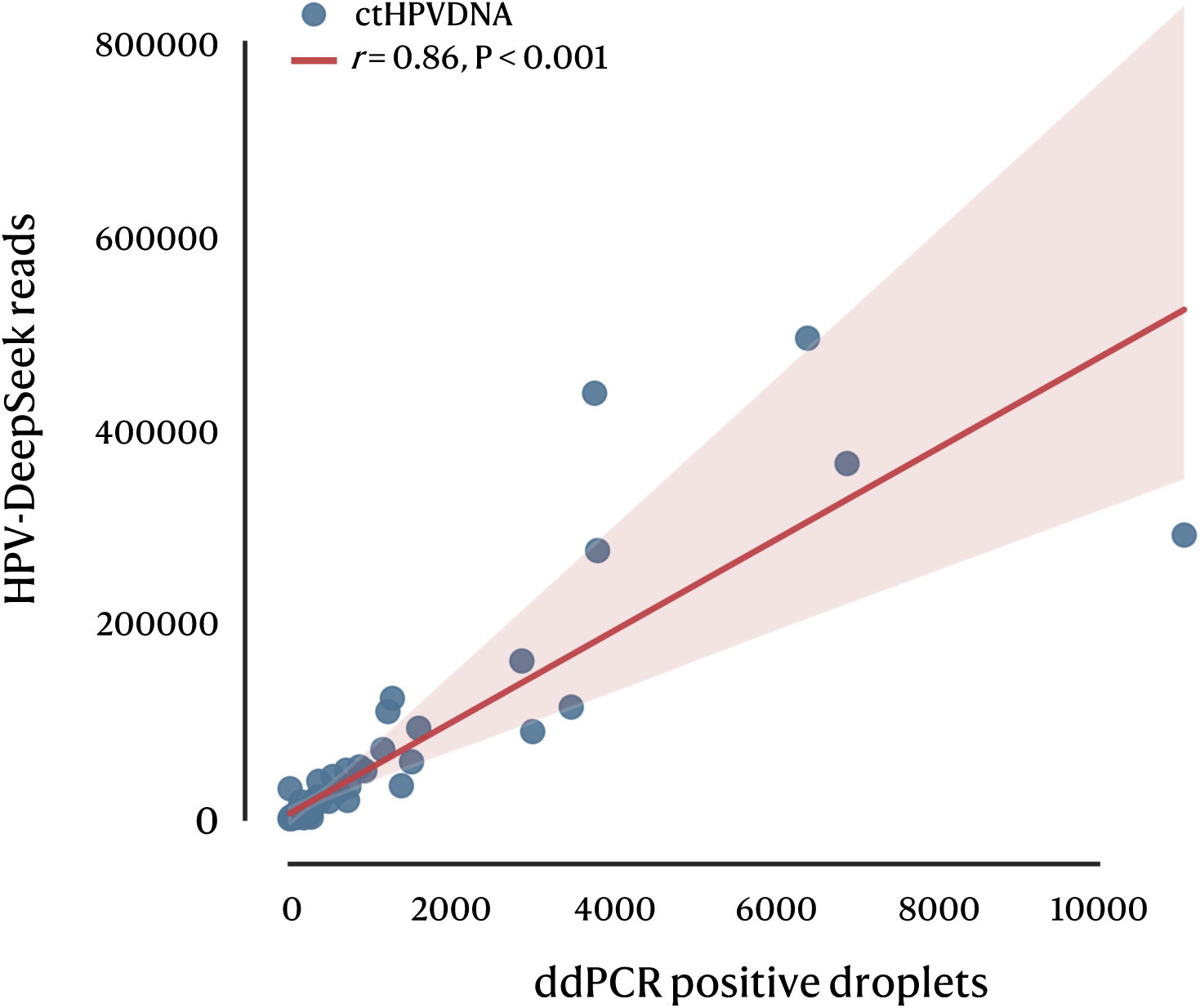
Linear correlation between ddPCR-positive droplets and HPV-DeepSeek reads. Each point represents an individual sample (blue). The red line shows the linear regression fit with 95% confidence interval (shaded). Pearson’s correlation coefficient was calculated on log-transformed values.

**Supplementary Figure 2.**
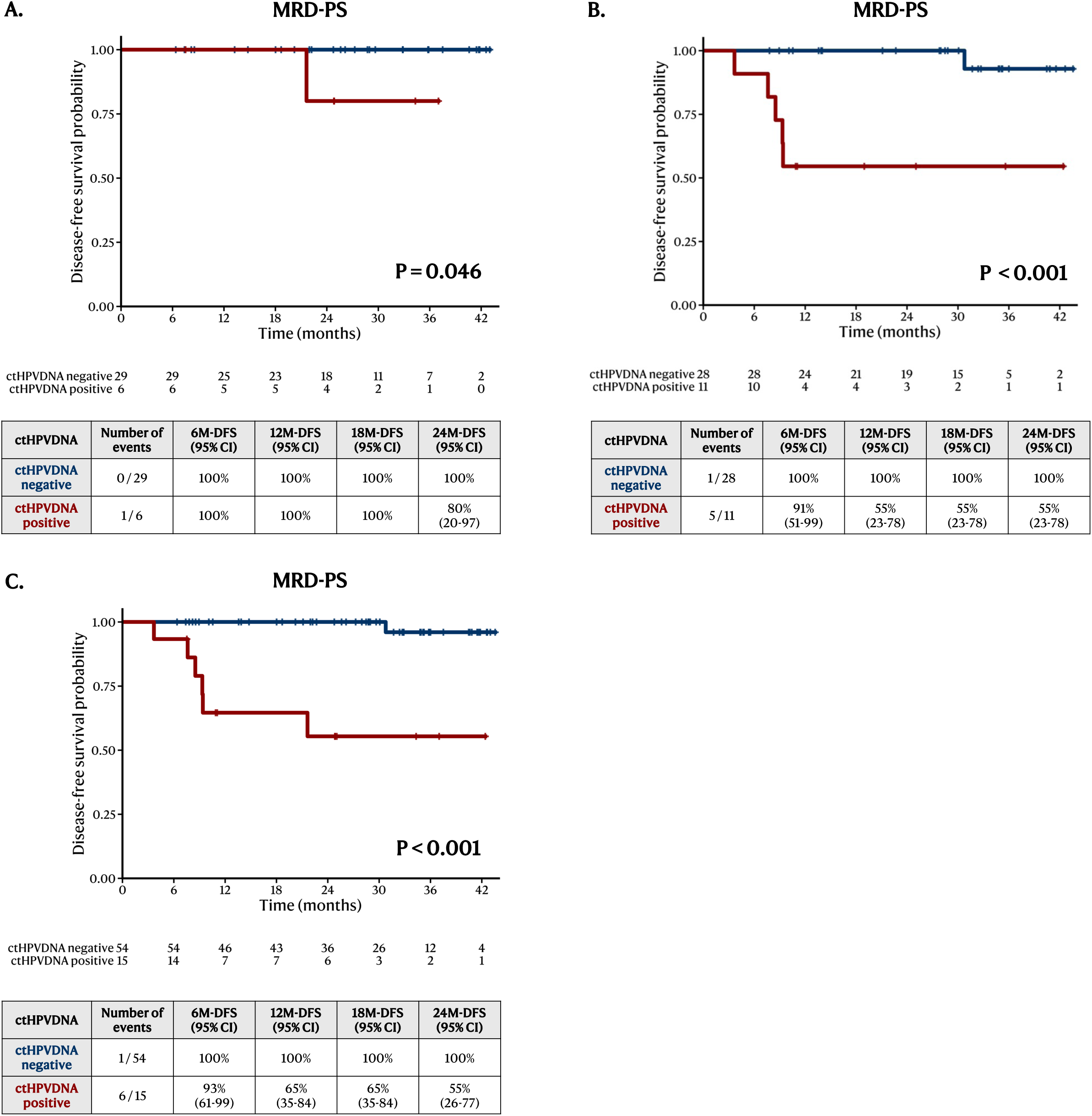
Association of MRD-PS status with recurrence by cancer stage and anatomical site. Kaplan–Meier DFS curves stratified by ctHPVDNA status during the MRD-PS window. (A) Early-stage disease (T0–T1, N0–N1) cohort. (B) Later-stage disease cohort. (C) Subgroup analysis restricted to oropharyngeal cancer cases. In each panel, patients with MRD-PS positivity (red lines) demonstrated significantly inferior DFS compared to MRD-PS negative patients (blue lines). Tables below each curve show number of events, number at risk, and disease-free survival rates at 6, 12, 18, and 24 months. P-values were calculated using the log-rank test.

**Supplementary Figure 3.**
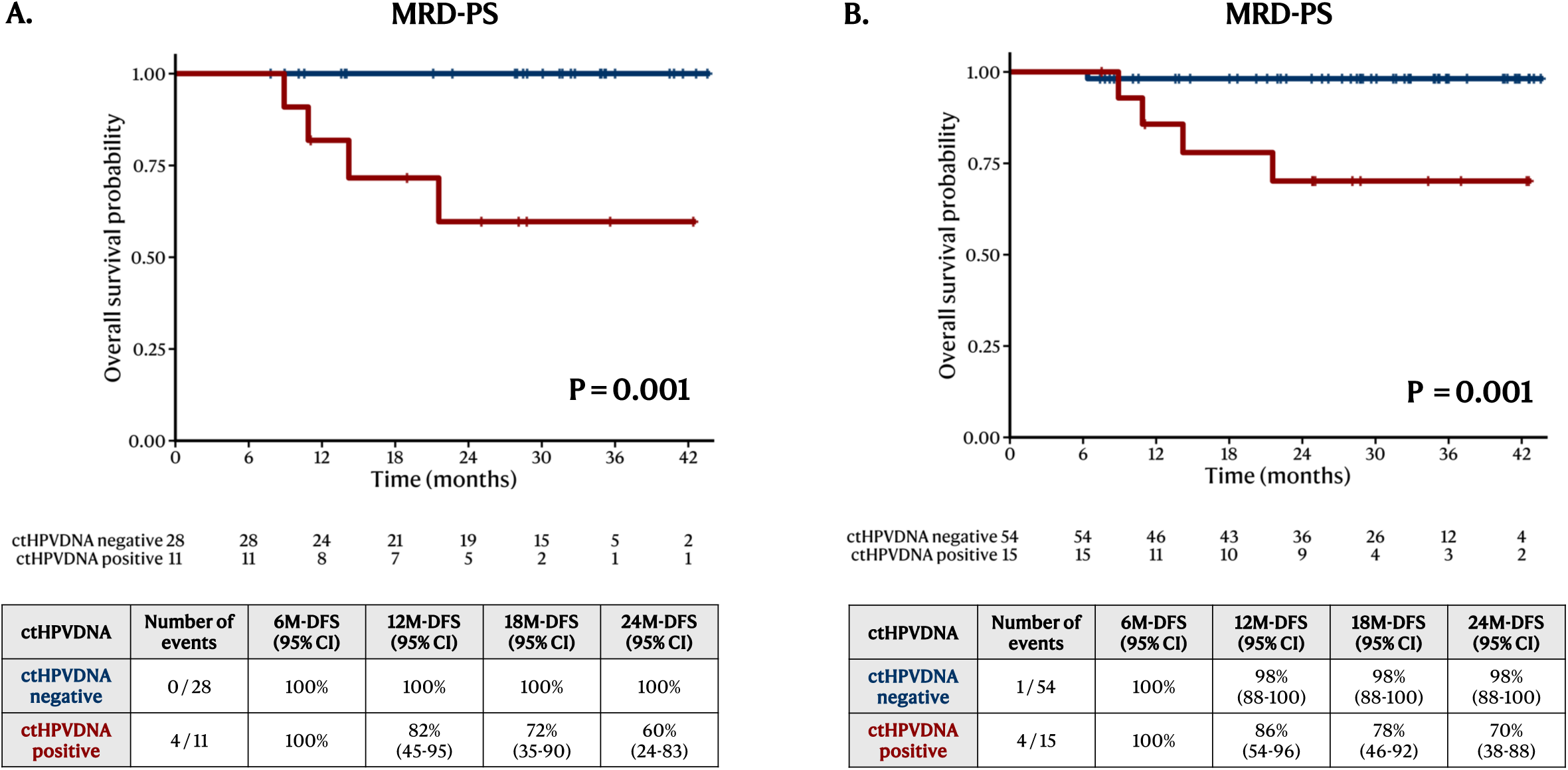
Association of MRD-PS status with death by cancer stage and anatomical site. Kaplan–Meier overall survival (OS) curves stratified by ctHPVDNA status during the MRD-PS window. (A) Later-stage disease cohort. (B) Subgroup analysis restricted to oropharyngeal cancer cases. MRD-PS positive patients (red lines) consistently exhibited significantly worse OS compared to MRD-PS negative patients (blue lines). Tables beneath each curve display number of events, number at risk, and overall survival rates at 6, 12, 18, and 24 months. P-values were calculated using the log-rank test.

**Supplementary Figure 4.**
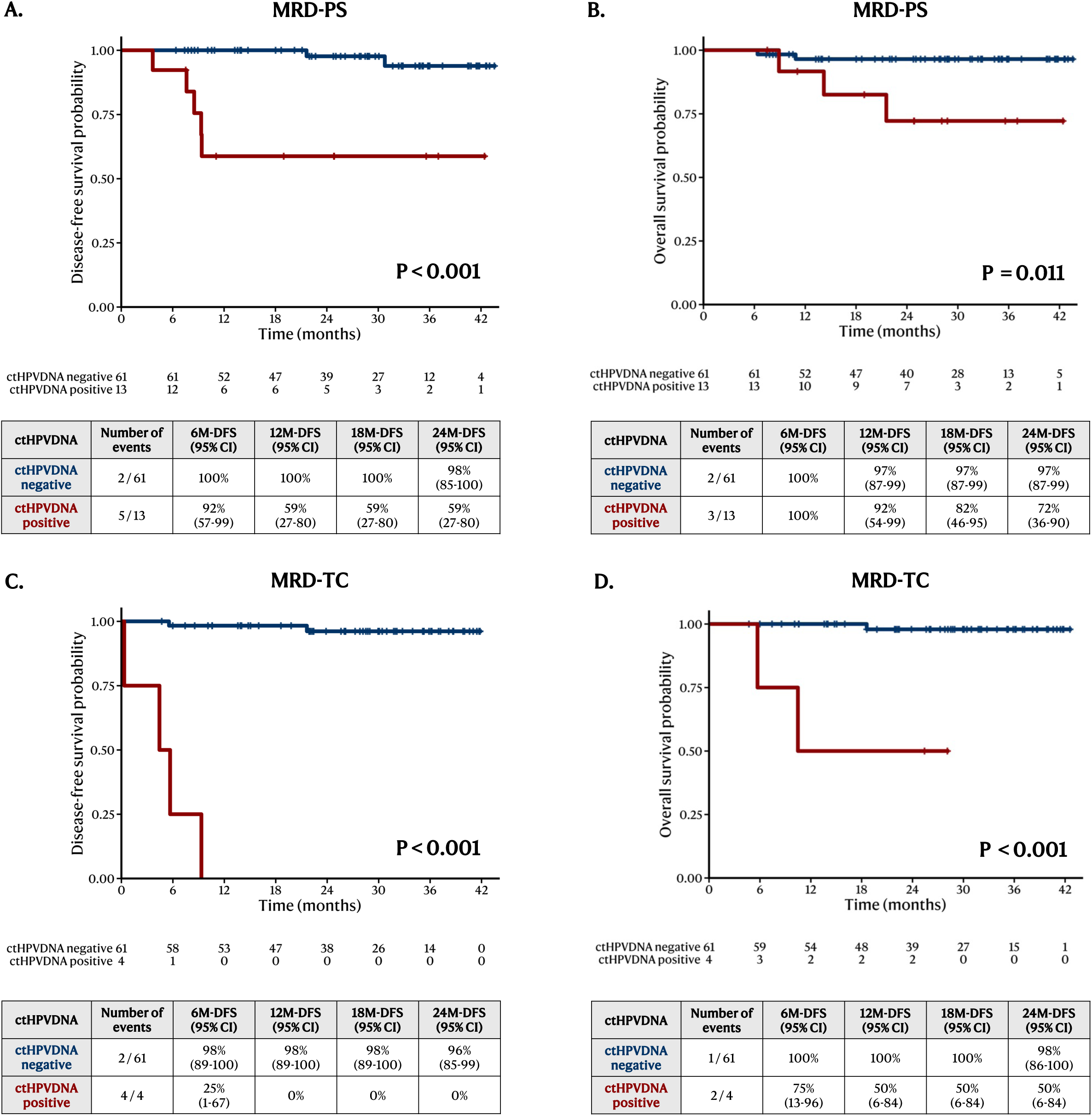
Prognostic value of single-time point ctHPVDNA detection. Kaplan–Meier survival curves evaluating disease-free survival (DFS) and overall survival (OS) stratified by single-timepoint ctHPVDNA status during MRD-PS and MRD-TC windows. (A) DFS based on single-time ctHPVDNA detection during the MRD-PS window. (B) OS during the MRD-PS window. (C) DFS during the MRD-TC window. (D) OS during the MRD-TC window. In each panel, ctHPVDNA-positive patients (red lines) had significantly worse survival outcomes compared to ctHPVDNA-negative patients (blue lines). Number at risk, number of events, and survival rates at 6, 12, 18, and 24 months are shown beneath each curve. P-values were calculated using the log-rank test.

**Supplementary Figure 5.**
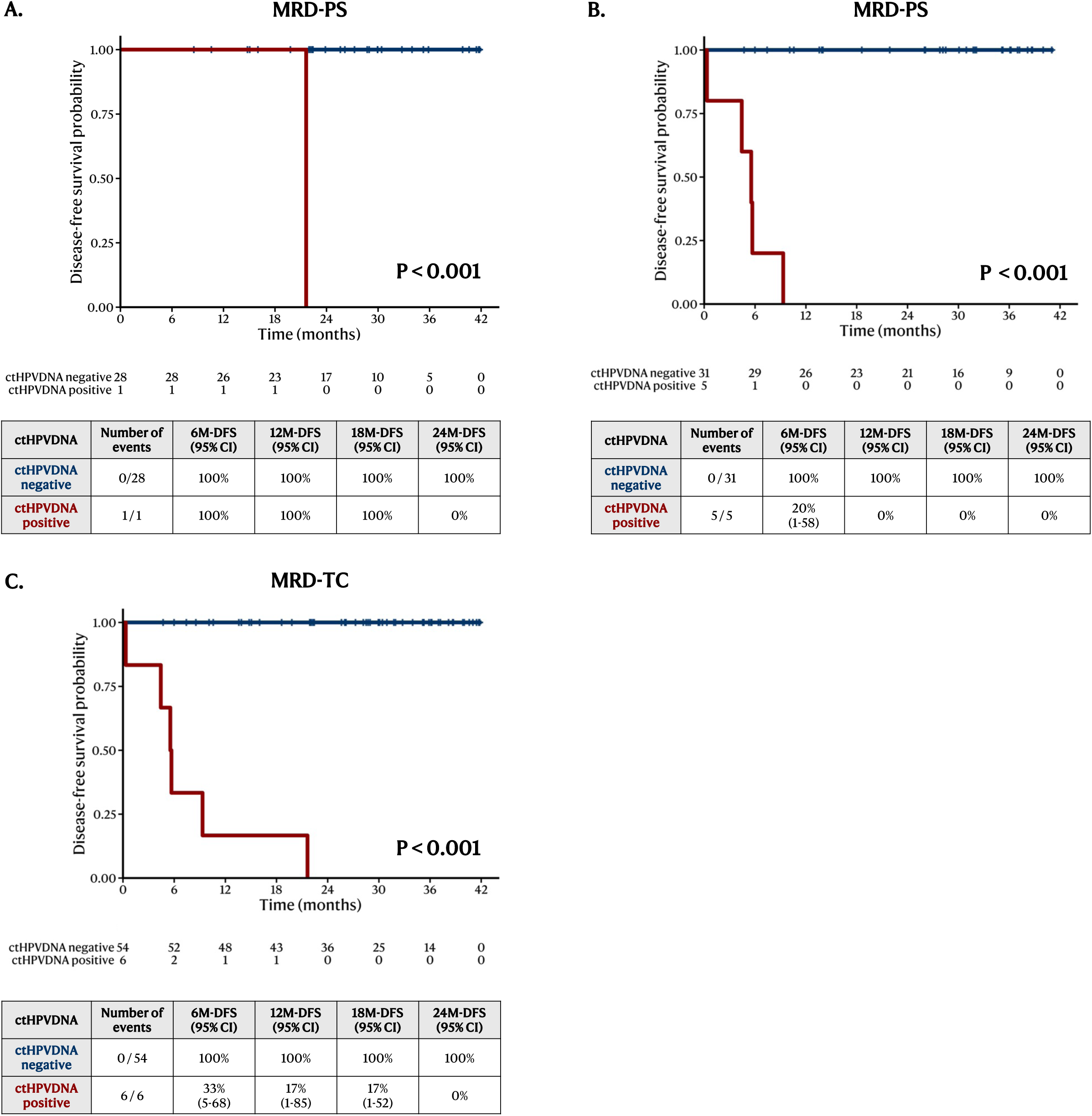
Association of MRD-TC status with recurrence by cancer stage and anatomical site. Kaplan–Meier disease-free survival (DFS) curves stratified by ctHPVDNA status during the MRD-TC window. (A) Early-stage disease (T0–T1, N0–N1) cohort. (B) Later-stage disease cohort. (C) Subgroup analysis restricted to oropharyngeal cancer cases. Across all groups, MRD-TC positivity (red lines) was strongly associated with significantly worse DFS compared to MRD-TC negativity (blue lines). Number at risk, number of events, and DFS rates at 6, 12, 18, and 24 months are shown below each curve. P-values were calculated using the log-rank test.

**Supplementary Figure 6.**
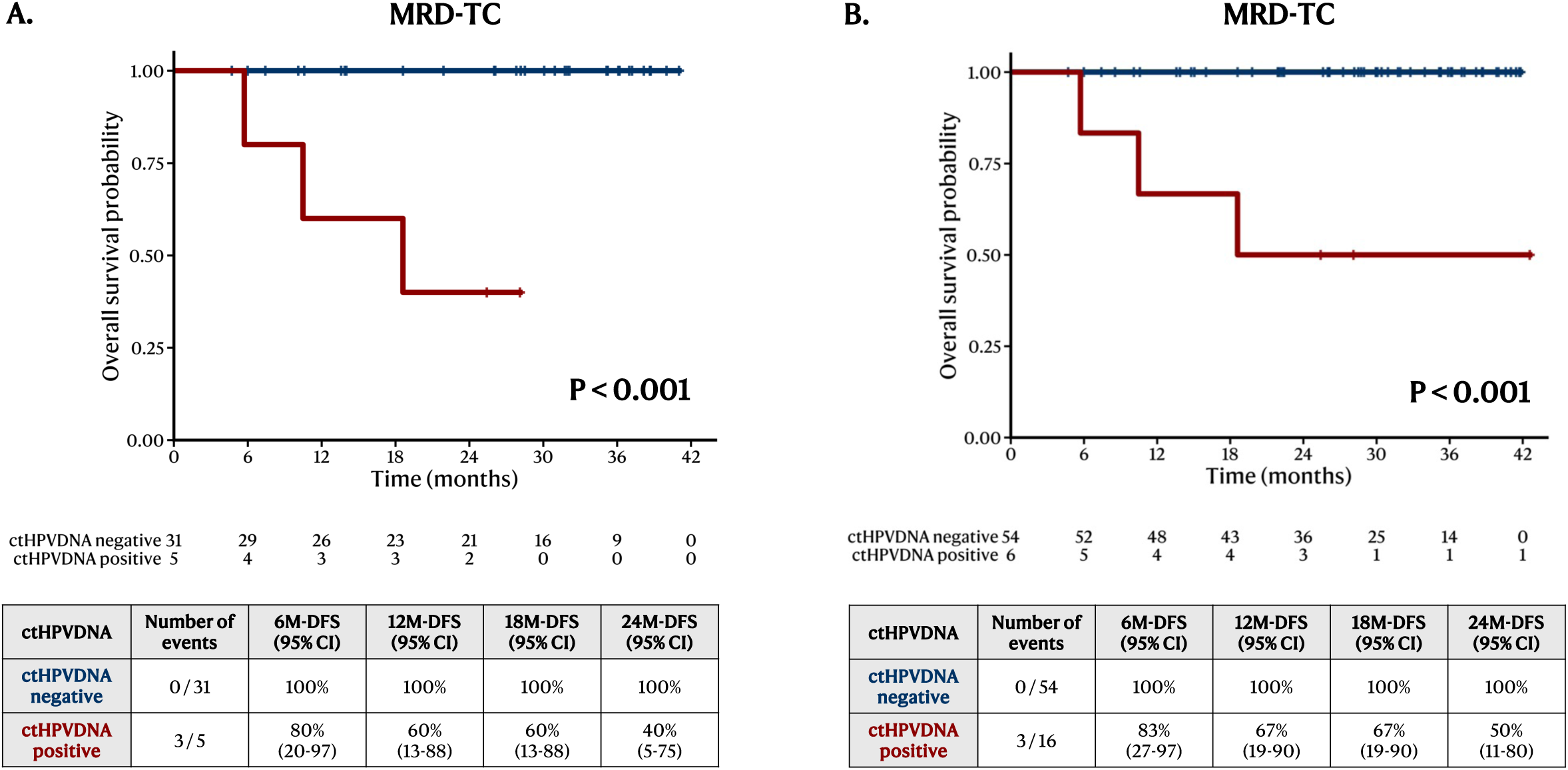
Association of MRD-TC status with death by cancer stage and anatomical site. Kaplan–Meier overall survival (OS) curves stratified by ctHPVDNA status during the MRD-TC window. (A) Later-stage disease cohort. (B) Subgroup analysis restricted to oropharyngeal cancer cases. In both groups, MRD-TC positivity (red lines) was significantly associated with inferior OS compared to MRD-TC negativity (blue lines). Tables below each curve display number at risk, number of events, and survival rates at 6, 12, 18, and 24 months. P-values were calculated using the log-rank test.

**Supplementary Figure 7.**
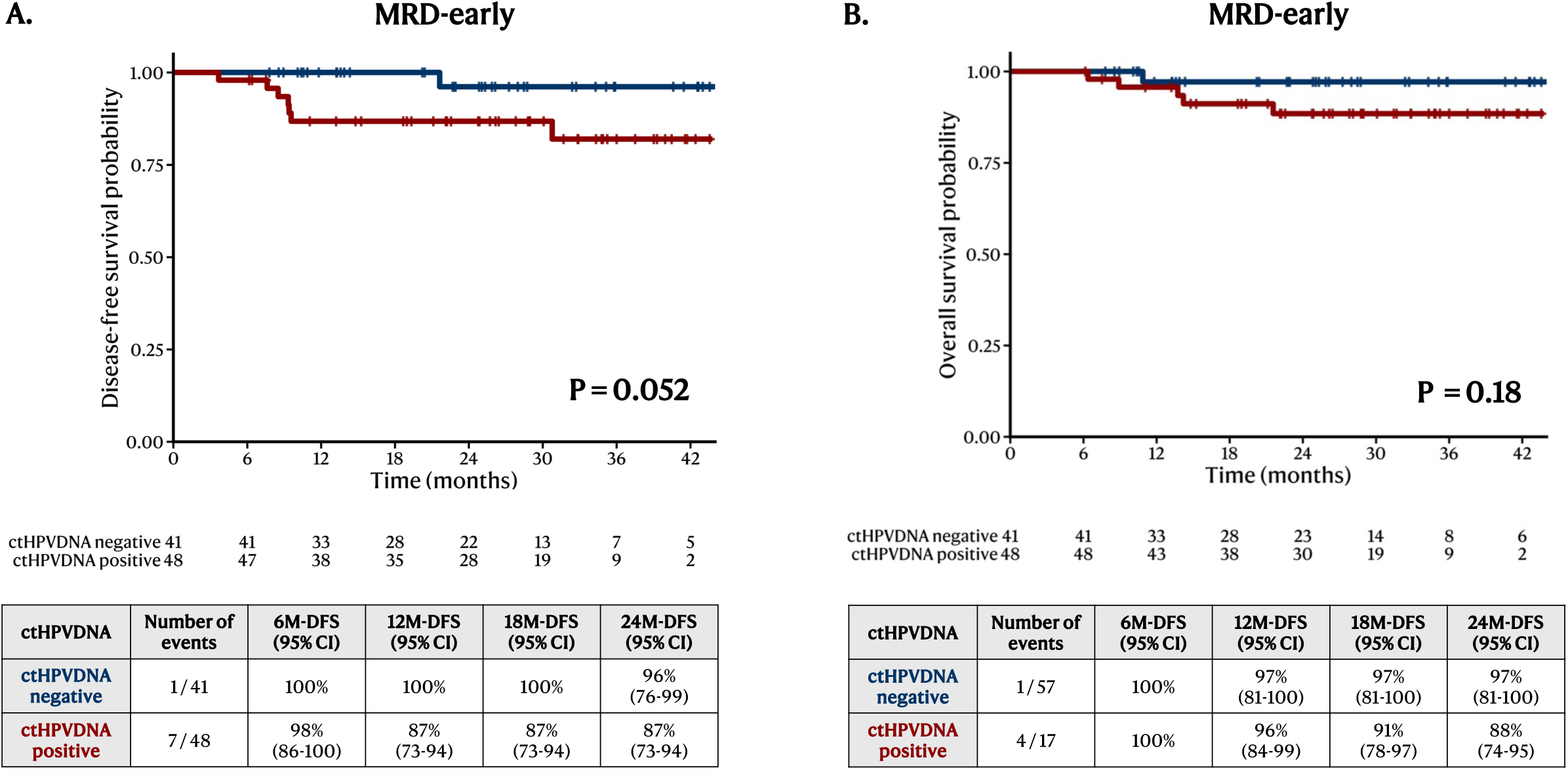
The prognostic value of ctHPVDNA immediately after surgery. Kaplan–Meier curves evaluating disease-free survival (DFS) and overall survival (OS) based on ctHPVDNA status in the MRD-early window (days 1–3 post-surgery). (A) DFS stratified by ctHPVDNA positivity (red line) versus negativity (blue line). (B) OS stratified by ctHPVDNA status. Tables below each curve display number at risk, number of events, and survival rates at 6, 12, 18, and 24 months. P-values were calculated using the log-rank test.

**Supplementary Figure 8.**
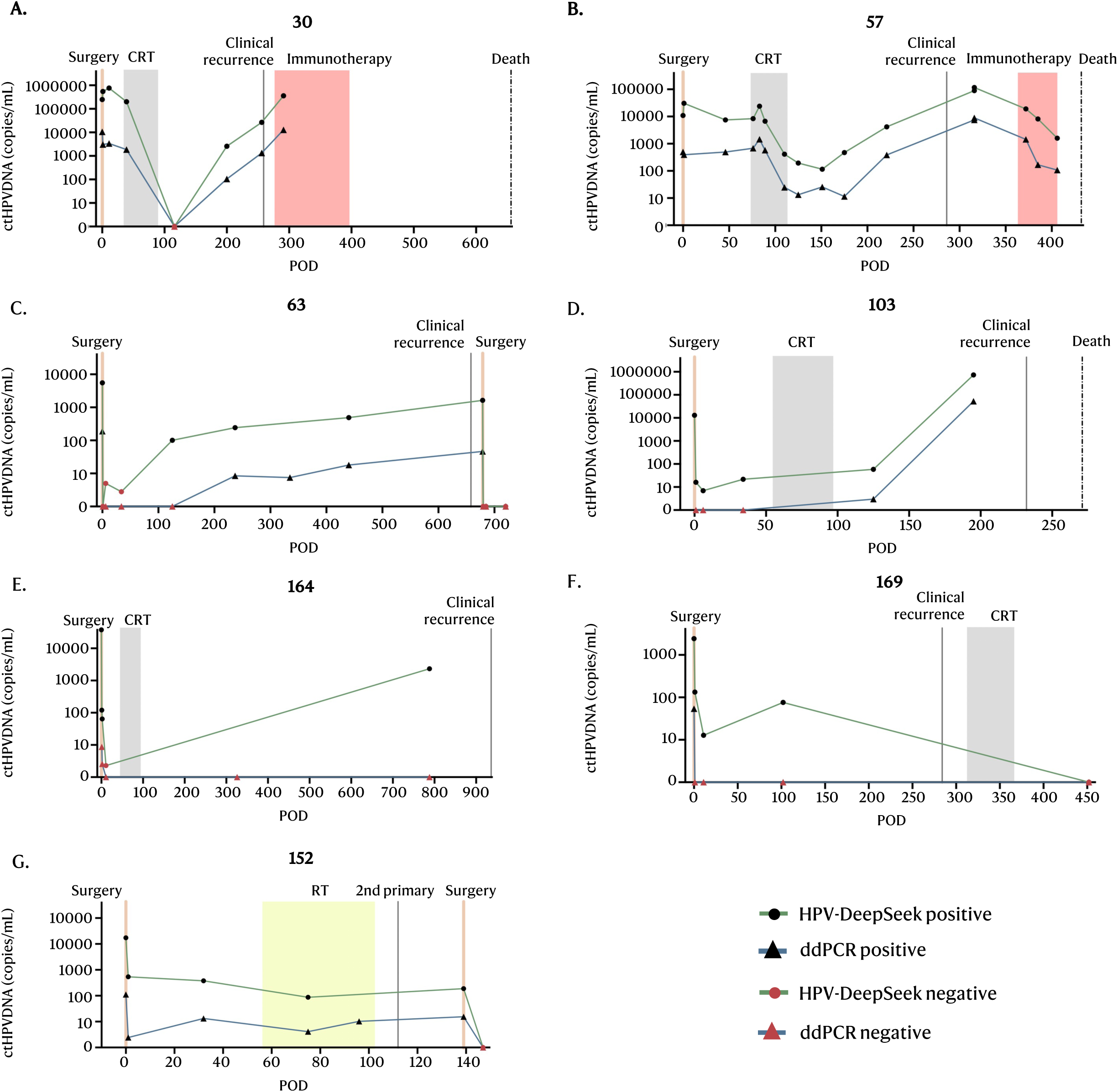
Longitudinal ctHPVDNA monitoring of recurrent cases. Line graphs showing the changes in ctHPVDNA levels over time for recurrent cases (A-F) and 2nd primary case (G) Each plot represents one patient’s ctHPVDNA trajectory as a function of time, with the sample collection timepoint in days after surgery on the x-axis and the ctHPVDNA levels on the y-axis (logarithmic scale). The green line represents the standardized ctHPVDNA value (*copies/ml) for HPV-DeepSeek, and the blue line for ddPCR. The markers are color-coded for positive and negative time points.

**Supplementary Table 1.**
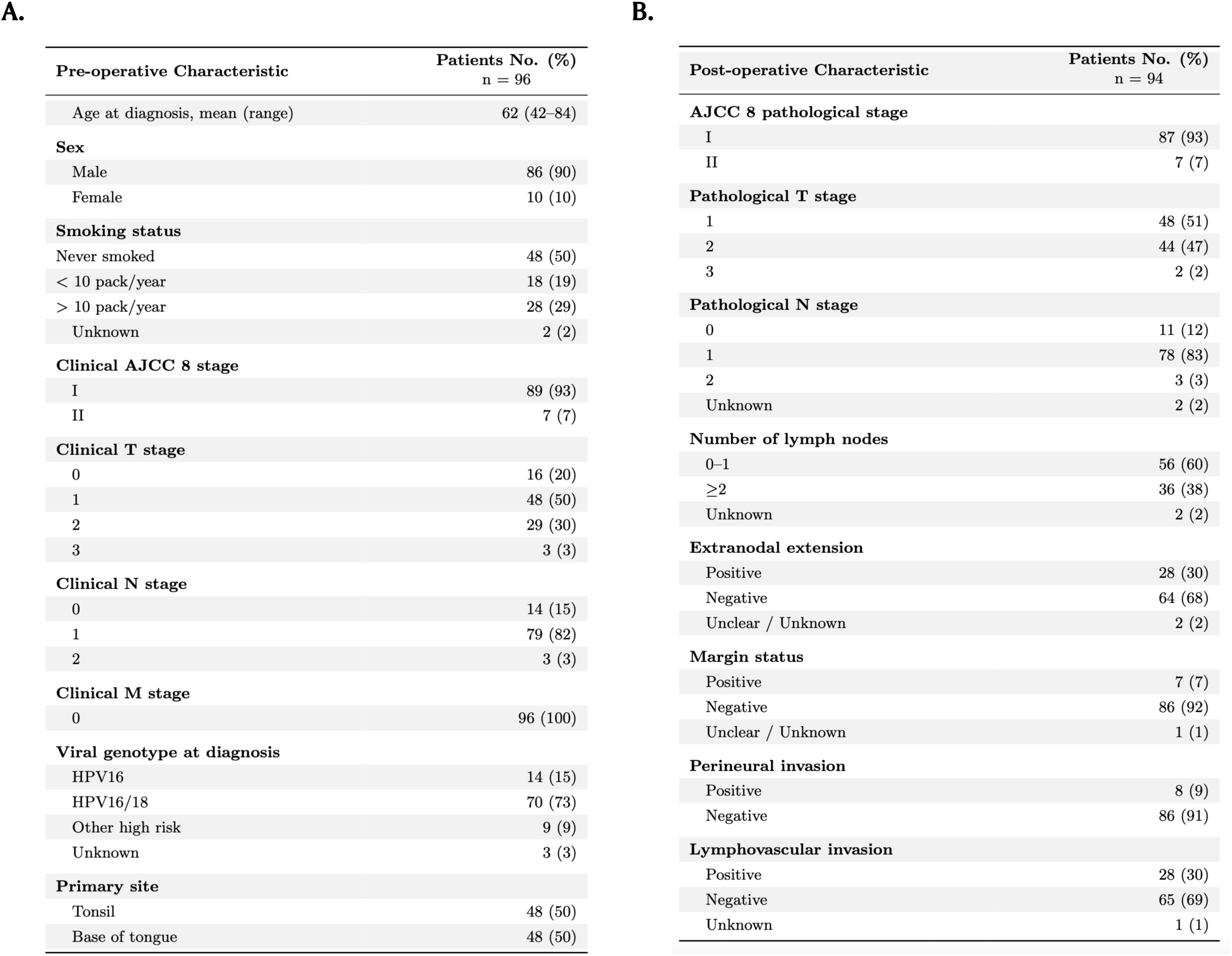
Patient demographics, staging, and clinicopathologic risk factors for oropharynx cancers. (A) Clinical staging and demographic characteristics of 96 HPV+ oropharynx cancer cases treated with surgery. (B) Pathological staging and clinicopathologic risk factors of 94 cases which were ctHPVDNA positive at the baseline and included in survival analysis.

**Supplementary Table 2.** Summary of ctHPVDNA Dynamics Across Clinical Windows and Patient Groups (A) Summary statistics for the absolute change in circulating tumor HPV DNA (Δ ctHPVDNA) between key clinical timepoints. For each transition window, median, mean, and standard deviation of Δ ctHPVDNA are shown across defined clinical groups, including MRD-negative, gradual, rapid, and MRD-positive patients. Kruskal-Wallis tests were used to compare distributions, with raw and FDR-adjusted p-values reported. (B) Summary of ctHPVDNA clearance slopes (log₁₀/day) across patient groups. For each transition window, the median, mean, standard deviation, and 95% confidence interval of the mean slope are reported for the cleared spontaneously versus the not cleared spontaneously groups. Group comparisons were made using Mann–Whitney U tests, with both raw and FDR-adjusted p-values shown.

| Window | Group | N | Median<br>($\Delta$ ct-HPVDNA) | Mean<br>($\Delta$ ct-HPVDNA) | SD<br>($\Delta$ ct-HPVDNA) | H-statistic | Raw *p* | FDR *p* |
| --- | --- | --- | --- | --- | --- | --- | --- | --- |
| Baseline →<br>MRD-early | MRD negative after<br>adjuvant treatment | 10 | −16,782 | −32,373.90 | 38,252.42 | 20.33 | < 0.001 | < 0.001 |
|  | Gradual | 22 | −17,329 | −67,363.32 | 124,127.85 |  |  |  |
|  | Rapid | 37 | −640 | −1,677.41 | 3,194.47 |  |  |  |
|  | Remained MRD<br>positive | 6 | −4,584 | −1,399.67 | 23,245.44 |  |  |  |
| MRD-early →<br>MRD-PS | MRD negative after<br>adjuvant treatment | 5 | −28 | −17.20 | 42.65 | 32.11 | < 0.001 | < 0.001 |
|  | Gradual | 22 | −75 | −574.36 | 2,137.02 |  |  |  |
|  | Rapid | 26 | −4 | −5.69 | 4.55 |  |  |  |
|  | Remained MRD<br>positive | 6 | −125 | −2,939.83 | 6,007.26 |  |  |  |
| MRD-PS →<br>MRD-TC | MRD negative after<br>adjuvant treatment | 7 | −48 | −994.14 | 2,539.62 | 9.95 | 0.019 | 0.019 |
|  | Gradual | 16 | −1 | −1.75 | 2.41 |  |  |  |
|  | Rapid | 9 | 0 | −0.22 | 1.56 |  |  |  |
|  | Remained MRD<br>positive | 4 | −11,252 | −104,131.50 | 193,678.35 |  |  |  |

| Window | Group | N | Median<br>slope<br>(log<sub>10</sub>/day) | Mean<br>slope<br>(log<sub>10</sub>/day) | SD of<br>slope | 95% CI of<br>mean slope | Raw *p* | FDR *p* |
| --- | --- | --- | --- | --- | --- | --- | --- | --- |
| Baseline →<br>MRD-early | Rapid/Gradual | 59 | −2.250 | −2.083 | 0.852 | −2.741 to −1.361 | 0.4189 | 0.4189 |
|  | Non-Rapid/Gradual | 16 | −1.818 | −1.796 | 1.211 | −2.574 to −1.072 |  |  |
| MRD-early →<br>Post-surgery | Rapid/Gradual | 48 | −0.063 | −0.098 | 0.106 | −0.125 to −0.026 | 0.0134 | 0.0403 |
|  | Non-Rapid/Gradual | 11 | −0.0049 | −0.0011 | 0.146 | −0.059 to<br>−0.0001 |  |  |
| Baseline →<br>Post-surgery | Rapid/Gradual | 58 | −0.251 | −0.280 | 0.213 | −0.395 to −0.092 | 0.3926 | 0.4189 |
|  | Non-Rapid/Gradual | 13 | −0.227 | −0.228 | 0.214 | −0.312 to −0.044 |  |  |

**Supplementary Table 3.** Impact of multi-vs single-timepoint sampling on recurrence and survival. Results were categorized based on two approaches within the MRD window: (1)’Single’, using only the earliest ctHPVDNA result, and (2)’Multiple’, using any positive sample to mean positive. DFS, OS, and their respective HRs are shown.

| MRD window<br>(group) | DFS<br>HR | DFS<br>MRD+ | DFS<br>MRD− | OS<br>HR | OS<br>MRD+ | OS<br>MRD− |
| --- | --- | --- | --- | --- | --- | --- |
| MRD-PS (multiple) | 25 | 60% | 100% | 14 | 73% | 98% |
| MRD-PS (single) | 16 | 59% | 98% | 7 | 72% | 97% |
| MRD-TC (multiple) | ∞ | 0% | 100% | ∞ | 50% | 100% |
| MRD-TC (single) | 116 | 0% | 96% | 38 | 50% | 98% |

**Supplementary Table 4.** Lead time for ctHPVDNA detection in recurrent cases by HPV-DeepSeek and ddPCR. Lead times by detection methods (HPV-DeepSeek vs ddPCR) and single-vs multi-time point samples. Green shading: ctHPVDNA was positive. Red shading: ctHPVDNA was negative. Longitudinal monitoring for patient 99 was not available. Patient 152 was a 2nd primary case, and thus, lead time was not calculated.

| Case | MRD after Treatment |  |  |  | MRD after Surgery |  |  |  | Recurrence | pT | pN | LN | LVI | PNI | ENE | Margins |
| --- | --- | --- | --- | --- | --- | --- | --- | --- | --- | --- | --- | --- | --- | --- | --- | --- |
|  | NGS<br>(single) | ddPCR<br>(single) | NGS<br>(multiple) | ddPCR<br>(multiple) | NGS<br>(single) | ddPCR<br>(single) | NGS<br>(multiple) | ddPCR<br>(multiple) |  |  |  |  |  |  |  |  |
| 30 | 0 | 0 | 59 | 59 | 248 | 248 | 248 | 248 | Distant | T2 | N1 | 4 | + | – | + | – |
| 57 | 161 | 161 | 161 | 161 | 240 | 240 | 240 | 240 | Multiple | T2 | N/A | N/A | + | + | N/A | + |
| 63 | 0 | 0 | 533 | 421 | 0 | 0 | 533 | 421 | Regional | T1 | N1 | 1 | – | – | – | – |
| 99 | N/A | N/A | N/A | N/A | N/A | N/A | N/A | N/A | Distant | T1 | N2 | 8 | + | – | + | – |
| 103 | 107 | 107 | 107 | 107 | 226 | 0 | 226 | 0 | Distant | T2 | N1 | 4 | – | – | + | – |
| 152 | N/A | N/A | N/A | N/A | N/A | N/A | N/A | N/A | 2nd primary | T2 | N1 | 3 | + | – | – | – |
| 164 | 148 | 0 | 148 | 0 | 0 | 0 | 0 | 0 | Distant | T2 | N1 | 3 | – | – | + | – |
| 169 | 273 | 0 | 273 | 0 | 273 | 0 | 273 | 0 | Regional | T2 | N1 | 1 | + | – | – | – |
| Sensitivity | 60% | 40% | 100% | 60% | 67% | 33% | 83% | 33% | – | – | – | – | – | – | – | – |
| Mean lead time | 115 | 45 | 214 | 125 | 165 | 81 | 253 | 152 | – | – | – | – | – | – | – | – |

